# Clinical Effect of Evolocumab treatment in coronary artery bypass surgery

**DOI:** 10.1101/2025.03.20.25324358

**Authors:** Giuseppe Nasso, Giuseppe Santarpino, Vincenzo Calabrese, Giovanni Taverna, Claudio Larosa, Isabella Rosa, Francesco Bartolomucci, Vincenzo Montemurro, Alessandro Fiorentino, Pasquale Mastroroberto, Felice Agrò, Giuseppe Speziale

## Abstract

**Background:** We demonstrated that Evolocumab treatment after coronary artery bypass graft surgery significantly reduced LDL-cholesterol and total cholesterol levels compared to statin treatment alone. However, in the short period following surgery, did not impact the reduction of major clinical events.

**Methods:** We collected dyslipidemic patients who underwent coronary artery bypass graft surgery following acute coronary syndrome or in an elective setting (254 patients). Since January 2020, we added to statin therapy with or without ezetimibe also an immediate postprocedural treatment with subcutaneous evolocumab, either 140 mg every 2 weeks; the previous period patients underwent a conventional treatment with statins. All patients were followed up clinically and laboratory, comparing the 2 groups (Standard vs Evolocumab).

**Results:** The variables were inserted into a Cox model, which found a significant impact for Group, Hypertension, EUROSCORE_II and previous stroke. In detail, the treatment with Evolocumab had a protective effect on the clinical events (recurrent angina, myocardial infarction, cerebrovascular events, need for coronary re-angiography, coronary artery re-angioplasty/re-bypass and cardiac death), with a HR of 0.38 (95% CI 0.15-0.99, p=0.047). Furthermore, cholesterol levels decreased more quickly over time in the evolocumab-treated group than in the conventional group, with an average difference of 27 mg/dl (p<0.01). Similarly, LDL decreased by approximately 30 mg/dl (p<0.001), triglycerides by approximately 18.77 mg/dl (p<0.001), while HDL increased by an average of 2.7 mg/dl more than in the conventional therapy (p<0.001).

**Conclusion:** The use of Evolocumab immediately after coronary artery bypass graft surgery, even in an urgent or elective setting, significantly reduced cholesterol levels compared to statin treatment alone. Moreover, this strategy in statin-resistant patients is also able to significantly reduce the major clinical cardiac events at follow-up.

## Introduction

Monoclonal antibodies that inhibit proprotein convertase subtilisin-kexin type 9 (PCSK9) are entering by right as a new class of drugs to lower LDL cholesterol levels (1); especially following an acute coronary syndrome (2). The basis of this effect is genetics: the carriage of PCSK9 loss-of-function alleles is linked to lower LDL cholesterol levels, reducing the risk of myocardial infarction (3,4).

In particular, the acute coronary syndromes (ACS) patients are at increased risk of recurrent ischemic events (5) and Evolocumab demonstrated to reduce major cardiovascular events in secondary prevention (6-9). Indeed, an early and strong strategy to lipid-lowering using PCSK9i in patients with ACS is safe and effective in clinical practice (10). However, the LDL-C-lowering efficacy of PCSK9 antibody treatment during the ACS in patients who underwent coronary artery bypass surgery (CABG) is unknown. Only in our previous study we analyzed and showed the short time effect of evolocumab in this cohort of patients (11). The CABG population is complicated to analyze and understand: ischemic and reperfusion myocardial injury is often reported in patients who underwent coronary artery bypass grafting (CABG) (12,13) despite substantial advancements in cardiopulmonary support devices and surgical techniques (14-16).

We recorded a significant reduction of the cholesterol and LDL in the patients who underwent evolocumab without a recorded clinical effect (11).

Also in elective CABG patients with hypercholesterolemia not statin responsive, we tried and demonstrated an immediate efficacy of evolocumab to ameliorate the laboratory (17). However, given the potential cardioprotective effects, we hypothesize that Evolucumab would give a beneficial clinical effect for patients undergoing cardiovascular surgery. Up to now, we are not able with the previous studies to demonstrate that. We are waiting for an ongoing study on the effect of PCSK9 inhibitors following bypass surgery (18) but in the present study we added the ACS patients who underwent CABG and the elective CABG patients for a longer follow-up focused on the clinical efficacy of the immediate treatment with evolucumab.

## Methods

From January 2017 to June 2023, at the cardiac surgery of Anthea Hospital GVM Care & Research Bari, 682 patients (568 men and 114 women) were operated on for isolated coronary surgery due to stable or atypical angina and coronary angiography study performed electively or patients with ST- and non-ST-segment elevation acute coronary syndrome (STE and NSTE-ACS) with symptom onset <72 h and clinical/hemodynamic stability. Of these elective patients, 230 were affected by dyslipidemia while being preoperatively treated with statins or not receiving cholesterol-lowering treatment at all. Among these dyslipidemic operated patients, 180 were visited one month after the operation and at the following follow-ups by the cardiac surgeon who evaluated the therapy taken by the patient and the laboratory tests, including the lipid profile.

Until December 2019, 100 patients with these characteristics (elective) were seen and of these 22 had an LDL-C concentration ≥70 mg/dL while on optimized lipid-lowering therapy, including a high-or moderate-intensity statin with or without ezetimibe and 78 patients with LDL-C concentration ≥70 mg/dL without statin therapy. Since January 2020, once Evolocumab was usable in patients with high LDL-C values, the next 80 patients with the same characteristics as the previous ones added statin therapy with or without ezetimibe (linked to the increase in CPK and transaminases at follow-up) also the treatment with subcutaneous Evolocumab either 140 mg every 2 weeks.

Speaking of the ACS patients (43 standard patients), they received the following protocol:

1. if without statins, they received a moderate-intensity statin (e.g. 40mg) from the cath lab table up to the 3-month follow-up;
2. on statin, they added Ezetimibe and a higher statin dosage (80 mg) from the cath lab table up to the 3-month follow-up;

Following the new scientific suggestions (29bis,30bis) from February 2023 in 31 patients with LDL > 100 mg/dL we changed our protocol to:

1. if without Statins, they received a moderate-intensity statin (e.g. 40mg) + Ezetimibe + PCSK9i (Evolocumab 140 mg every 2 weeks) from the cath lab table up to the 3-month follow-up;
2. on statin, they added Ezetimibe and a higher Statin dosage (80 mg) + PCSK9i (Evolocumab 140 mg every 2 weeks) from the cath lab table up to the 3 months follow up; The PCSK9i was administered at baseline “as early as possible”, usually before the coronary angiography.

These patients were followed on a clinical and laboratory level and the data were compared between the 2 groups (evolocumab treated versus conventional treatment), in particular the results were compared in terms of the ability to have an effect on the laboratory values of LDL-C and clinically: risks of recurrent angina, myocardial infarction, cerebrovascular events, need for coronary re-angiography, coronary artery re-angioplasty/re-bypass and cardiac death were analyzed.

The patients provided written informed consent before participating in the study. The corresponding author had full access to all the data in the study and takes responsibility for the integrity of the data and the accuracy of the data analysis.

### Data and Material Availability

The data used and analyzed in this study are available upon reasonable request from the corresponding author. The analysis was conducted in accordance with the transparency and openness (TOP) guidelines of AHA journals. All methodological details, including patient selection criteria, treatment procedures, and statistical analyses, are described in the manuscript. Any additional data or supplementary materials can be provided upon request, in compliance with ethical regulations and patient privacy policies.

Local Ethical Committee approved the study with two different protocols for elective (Prot. N. 001.06.96. Study number 7815) and “acute coronary syndrome” patients (Prot. N. 22.01.24. Study number 7809).

### Statistical Analysis

Data were described as mean ± standard deviation, median and interquartile range, or proportion, as appropriate. The distribution of variables was investigated by the Kolmogorov-Smirnov test followed by graphic evaluation. No missing data was retrieved in the database. The sample was divided into two groups based on Evolocumab therapy. 111 patients took Evolocumab and 143 were allocated in the control group. The comparisons between the two groups were computed using student T-test, the Mann-Whitney test the Pearson Chi-square test, as appropriate. A machine learning Cox regression method with ridge regularization was applied to detect confounding variables. Variables with impact above the median were included in the multivariate Cox analysis. Furthermore, generalized linear models for repeated measures were used to detect the differences in total cholesterol, LDL, HDL, triglycerides and ALT slopes in time between the two groups.

## Results

254 patients were enrolled in the analysis. Patients were aged 68 years [63-73], males were 190 (75%), with a BMI of 27 [24-29] kg/m2. 126 (50%) had arterial hypertension, 70 (28%) had diabetes, 73 (29%) had smoking habits. 39 patients manifested CV events (15%), 7 (6%) in the Evolocumab group and 32 (22%) in the control group (p<0.001). On the basis of this difference in the event incidence, a sample of 68 patients for each group was enough to reach the statistical power of 80%. (Figure 1).

**Figure 1.**
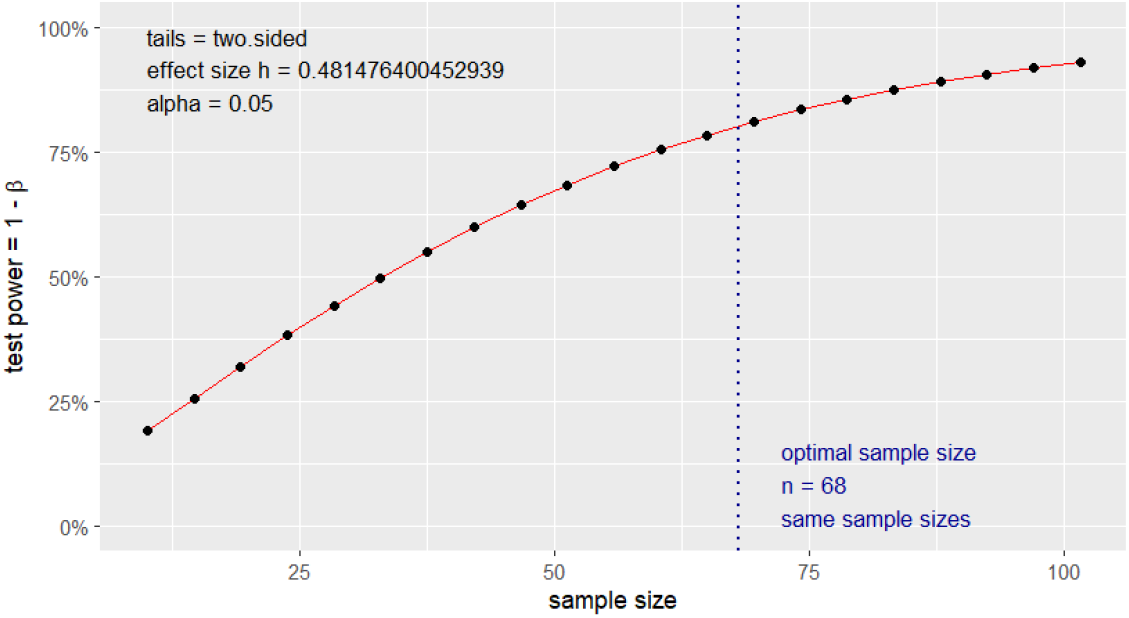
Difference of proportion power calculation for binomial distribution (arcsine transformation)

Table 1 shows the preoperative characteristics of the 2 groups (conventional and evolocumab treated). The two groups differed at the baseline for age, BMI, arterial hypertension, HDL, triglycerides and ALT. In detail, evolocumab group was older, with a lower BMI, lower LDL and Higher ALT. Using machine learning methods, we identified which variables are most associated with the event and we took those with an impact higher than the median (table 2). Variables with an impact coefficient on event above the median (0.00532), detected by Ridge Cox regression, were BMI, diabetes, smoking habits, previous IMA, previous coronary artery bypass surgery, previous stroke, vascular diseases, malignancies, EUROSCORE II, arterial hypertension, number of bypasses, sex and Evolocumab use (table 2). These variables have been included in the multivariate Cox regression model, which showed a significant impact for Evolocumab use, arterial hypertension, EUROSCORE II, and history of stroke. In detail, EVO reduced the risk of events, with an HR of 0.39 (95% CI 0.15-0.99, p=0.047). Harrel’s C index of this model was 0.7338 (Table 3). Furthermore, cholesterol levels decreased more quickly over time in the evolocumab-treated group than in the conventional group, with an average difference of 27 mg/dl (p<0.01). Similarly, LDL decreased by approximately 30 mg/dl (p<0.001), triglycerides by approximately 18.77 mg/dl (p<0.001), while HDL increased by an average of 2.7 mg/dl more than in the conventional therapy (p<0.001).

**Table 1.**
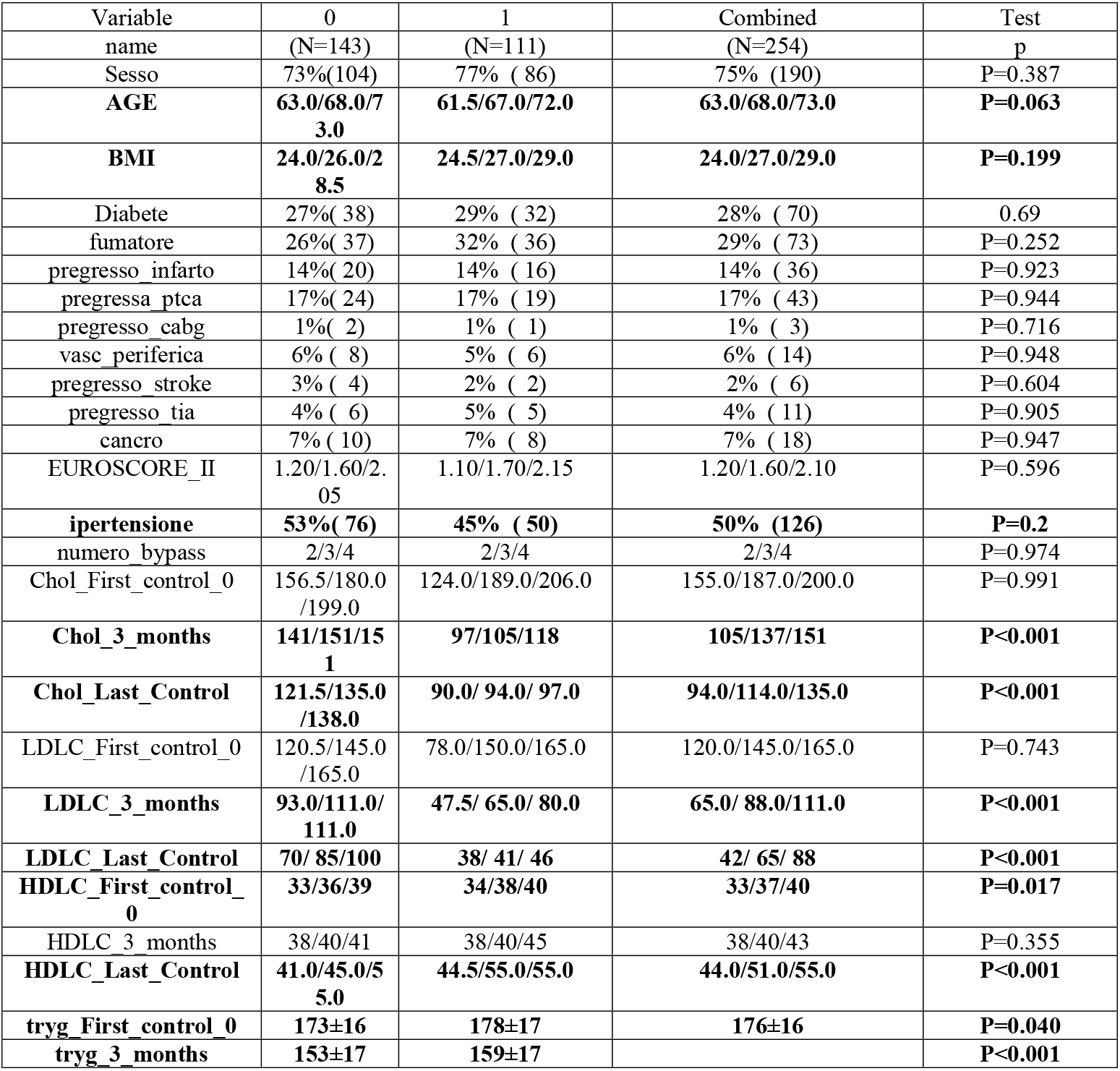
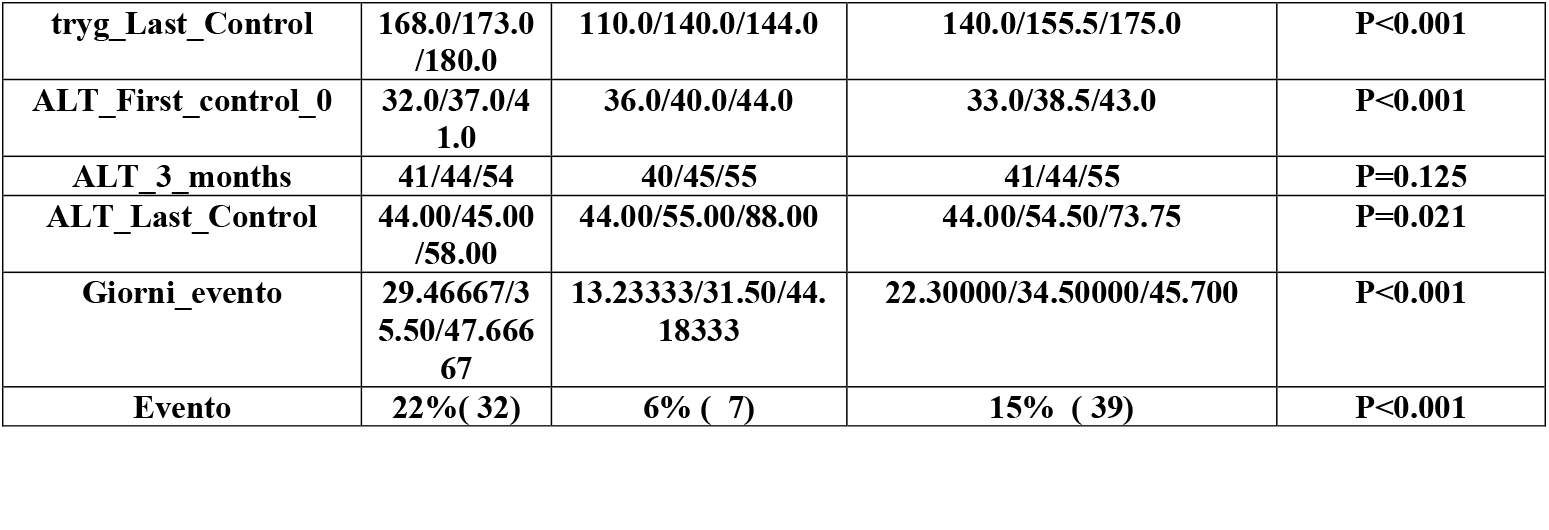
Preoperative characteristics.

**Table 2.** Impact of clinical variables on event risk.

| Variable | Impact<br>coefficient |
| --- | --- |
| AGE | 0.0040502744 |
| **BMI** | **0.0072390223** |
| **Diabete** | **0.0066197353** |
| **fumatore** | **0.0131524709** |
| **pregresso_infarto** | **0.0814036518** |
| pregressa_ptca | 0.0006076540 |
| **pregresso_cabg** | **0.1324897847** |
| **vasc_periferica** | **0.0563306224** |
| **pregresso_stroke** | **0.4619209711** |
| **pregresso_tia** | **0.0456872396** |
| **cancro** | **0.0300703973** |
| **EUROSCORE II** | **0.1120751035** |
| **ipertensione** | **0.0672182412** |
| **numero_bypass** | **0.0264609981** |
| Chol_First_control_0 | 0.0014086978 |
| Chol_3_months | 0.0003944460 |
| Chol_Last_Control | 0.0001661643 |
| LDLC_First_control_0 | 0.0014562843 |
| LDLC_3_months | 0.0005380014 |
| LDLC_Last_Control | 0.0015931742 |
| HDLC_First_control_0 | 0.0045593010 |
| HDLC_3_months | 0.0035853646 |
| HDLC_Last_Control | 0.0072897141 |
| tryg_First_control_0 | 0.0005259851 |
| tryg_3_months | 0.0004993233 |
| tryg_Last_Control | 0.0008237059 |
| ALT_First_control_0 | 0.0042528122 |
| **ALT_3_months** | **0.0053184538** |
| ALT_Last_Control | 0.0015605045 |
| **GRUPPO2** | **0.0613137202** |
| Sesso | 0.1041491524 |

**Table 3.** Multivariate Cox regression analysis of clinical variables associated with events.

| Variable | HR | 95% CI | p |
| --- | --- | --- | --- |
| Evolocumab | 0.39 | 0.15-0.99 | 0.047 |
| Sex | 0.73 | 0.51-1.07 | 0.409 |
| ALT 3 MONTH | 1.04 | 0.93-1.01 | 0.057 |
| N BYPASS | 0.99 | 0.70-1.41 | 0.969 |
| Arterial hypertension | 0.45 | 0.21-0.98 | 0.044 |
| EUROSCORE II | 2.49 | 1.14-5.40 | 0.021 |
| Malignances | 1.45 | 0.21-9.69 | 0.702 |
| Pregresso stroke | 13.46 | 1.71-36.2 | 0.001 |
| Vascular disease | 0.14 | 0.02-1.71 | 0.128 |
| Pregresso cabg | 0.26 | 0.03-2.59 | 0.547 |
| Pregresso IMA | 0.54 | 0.14-2.06 | 0.368 |
| Smoke | 1.44 | 0.56-3.67 | 0.452 |
| diabetes | 0.96 | 0.42-2.18 | 0.929 |
| BMI | 0.96 | 0.85-1.08 | 0.520 |

## Discussion

We previously demonstrated that evolocumab is able to reduce the cholesterol-total and LDL after cardiac surgery in elective patients (17) and in patients who underwent acute coronary syndrome (11). However, given its potential cardioprotective effects, it can be speculated that Evolocumab would be beneficial for patients undergoing cardiovascular surgery. In the present paper, with a longer follow-up, the patients who underwent evolocumab treatment suffered less cardiac events (combined risks of recurrent angina, myocardial infarction, cerebrovascular events, need for coronary re-angiography, coronary artery re-angioplasty/re-bypass and cardiac death).

The efficacy of PCSK9 inhibitors in patients undergoing cardiovascular surgery has not been demonstrated. We present our preliminary results to investigate the efficacy of a PCSK9 inhibitor (evolocumab) in preventing ischemic and reperfusion myocardial injury in multivessel coronary disease patients undergoing elective CABG surgery.

Due to these clinical consequences, the results of our study confirm the Fourier Trial (7) in the cardiac surgery field: they found that the addition of evolocumab to statin therapy significantly reduced the risk of cardiovascular events, with a 15% reduction in the risk of cardiovascular death, myocardial infarction, stroke, hospitalization for unstable angina, or coronary revascularization. In the same way, we recorded 32 events in the conventional therapy patients (22%) and 7 events in the evolocumab-treated patients (6.3%); a reduction of 15.7%.

These positive long-term effects of the addition of Evolocumab are, however, probably still only partially known, given that the long-term results of the therapy in patients at cardiovascular risk have also been published, demonstrating that the long-term LDL-C lowering with Evolocumab was associated with persistently low rates of adverse events that did not exceed those observed in the original placebo arm (19). Therefore, also in support of our study, it appears important to underline the early intake of the drug itself as proposed by us in our approach immediately, i.e. one month, after the surgical revascularization procedure. On the other hand, an early start is a hypothesis on which our study is based and has had clear bibliographical support for several years. In fact, a delay between the onset of LDL cholesterol lowering and the emergence of the full clinical benefit of the intervention in terms of clinical risk reduction has been well documented in many trials (7,20-23). The benefit of decreasing LDL cholesterol levels therefore remains indisputable, given that even studies prior to the use of evolocumab showed significant reductions in major cardiovascular events using statins alone in the Pravastatin or Atorvastatin Evaluation and Infection Therapy–Thrombolysis in Myocardial Infarction (PROVE IT–TIMI) and Treating to New Targets (TNT) trials (24, 25).

We have to highlight that our study is the first clinical and laboratory analysis of the evolocumab initiated in-hospital in patients presenting with STE- and NSTE-ACS underwent urgent surgical revascularization or elective CABG; and we recorded a difference in the cardioprotective effect associated with a substantially greater reduction in LDL-C levels. Our results build upon previous studies that investigated Evolocumab in ACS (26), patients with statin intolerance (27) or familial hypercholesterolemia (28), and patients with stable manifestations of ischemic cardiomyopathy (29, 30). The ODYSSEY OUTCOMES trial assessed the cardiovascular effects of alirocumab in patients at least 1 month (median 2.6 months) after an ACS (31). In our study we were not able to demonstrate an immediate clinical effect (17); but our actually longer follow-up is able to obtain the goal. Also following a longer follow-up, the incidence of adverse events was overall similar between groups. So, we confirm that the results are consistent with safety and tolerability data from previous studies with evolocumab in more stable clinical settings.

The limitation of our study is the summary of two different populations, but our two previous studies (11, 17) are the only publications on evolocumab in cardiac surgery in urgent and elective fields. Obviously, a prospective randomized trial is mandatory to confirm our results.

In conclusion, **i**n patients presenting with ACS or elective coronary disease who underwent CABG, Evolocumab initiated “as soon as possible” with high-intensity statin therapy and ezetimibe was well tolerated and resulted in a substantial reduction in LDL-C levels compared with only statin + ezetimibe. These laboratory results are associated with a reduction of the major clinical cardiac events at follow-up.

## Non-standard Abbreviations and Acronyms

ACS: Acute Coronary Syndrome
CABG: Coronary Artery Bypass Grafting
CPK: Creatine Phosphokinase
EUROSCORE II: European System for Cardiac Operative Risk Evaluation II
IMA: Acute Myocardial Infarction
LDL-C: Low-Density Lipoprotein Cholesterol
NSTE-ACS: Non-ST-Segment Elevation Acute Coronary Syndrome
PCSK9: Proprotein Convertase Subtilisin/Kexin Type 9
PCSK9i: PCSK9 Inhibitor
STEMI: ST-Elevation Myocardial Infarction
STE-ACS: ST-Segment Elevation Acute Coronary Syndrome

## Acknowledgements

We would like to thank all the authors for their valuable contributions and participation in the study, and we thank Jlenia D’Agnano, Veronica D’Anna and Valeria Cosco for manuscript review and editorial assistance.

